# Sex Disparity in Referral for Catheter Ablation for Atrial Fibrillation at a Single Integrated Health System

**DOI:** 10.1101/2024.01.29.24301970

**Authors:** Arati A. Gangadharan, Lutfiyya N. Muhammad, Jing Song, Bradley Knight, Rod Passman

**Author notes:** Correspondence to: Arati A. Gangadharan, MD, Northwestern Memorial Hospital, 676 N. St. Clair Street, Suite 600, Chicago, IL 60611.

## Abstract

**Background:** Catheter ablation for atrial fibrillation (AFCA) is indicated for select patients with atrial fibrillation (AF) and has been shown to reduce AF burden and improve quality of life. Earlier studies demonstrated that women are less likely to undergo AFCA despite having more AF symptoms. We investigated whether an association exists between referral patterns and this sex disparity.

**Methods and Results:** A retrospective cohort study was conducted of outpatients with newly diagnosed AF using the electronic medical record at Northwestern. Of 5,445 patients analyzed, 2,108 were women, and 3,337 were men. Characteristics including race, insurance status, comorbidities, and prior AF treatment were compared by sex. Logistic regression models adjusted for socioeconomic and clinical factors were constructed to determine associations between sex and binary dependent variables including referrals to and visits with general cardiology and electrophysiology (EP) and utilization of AFCA. There were no significant differences in odds of referral to (aOR, 1.17 [0.92-1.48], P=0.20) or visits with (aOR, 1.03 [0.82-1.30], P=0.79) general cardiologists between women and men. There were no significant differences in odds of referral to (aOR, 0.83 [0.68-1.01], P=0.06) or visits with (aOR, 0.87 [0.72-1.05], P=0.15) electrophysiologists between women and men. Finally, no significant difference was found in likelihood to undergo AFCA between women and men (aOR, 1.08 [0.83-1.40], P=0.56).

**Conclusions:** Our study demonstrates no significant differences in referral patterns to specialists or rates of AFCA between women and men. Encouraging equitable referral to specialists and access to AFCA is essential in ensuring appropriate care for all patients.

## Introduction

Atrial fibrillation (AF) is the most common sustained cardiac arrhythmia, estimated to affect millions of Americans every year. Its prevalence has increased three-fold in the last fifty years.^1^ As the proportion of elderly individuals continues to increase in our population, it is expected that the number of patients with AF will also increase 2.5-fold over the next fifty years leading to an estimated AF burden of 12.1 million Americans in 2030.^2,3^ Given that AF is considered a leading preventable cause of ischemic stroke and increases the risk of heart failure, dementia, and premature death, prioritizing timely and equitable management of AF has risen to the forefront of scholarship.^1^

Epidemiological investigation of AF has uncovered sex-based differences in AF presentation and management. Incident AF tends to be higher in men than women; however, given that women live longer than men, AF is more common in women above the age of 75 than their male counterparts.^4-6^ Women experience a more pronounced symptom burden of AF than men including palpitations, chest pain, dizziness, fatigue, and syncope.^7,8^ Additionally, women with AF suffer from greater functional limitations and a worse quality of life, which are products of greater symptoms, inability to engage in daily activities, and concern over AF management driving them to seek medical care more frequently.^6, 9-12^ Despite women with AF being more symptomatic and more likely to seek medical care, research has found that they are less likely to receive specialist care and rhythm control therapy.^6^

Catheter ablation for atrial fibrillation (AFCA) has been demonstrated to improve quality of life and may be pursued as an appropriate course of action for symptomatic AF prior to or following a rhythm control strategy with antiarrhythmic agents.^13^ However, prior studies have demonstrated that women are less likely to receive AFCA.^1, 12, 14, 20-23^ We investigated whether an association exists between referral patterns and this previously observed sex disparity in use of ablation.

## Methods

A retrospective cohort study was performed on data obtained from the Northwestern Medicine Enterprise Data Warehouse, a collection of clinical information sourced from the electronic medical record (EMR) of the Northwestern Medicine system. Data extraction was restricted to those patients diagnosed with AF at an outpatient encounter in the Central region, a geographic restriction encompassing a racially diverse downtown territory of the greater Chicago area. This study was approved by the Northwestern University Institutional Review Board. The requirement for informed consent was waived.

Inclusion criteria included age ≥18 years diagnosed with AF in internal medicine/primary care clinic (IM/PC), general cardiology clinic, or heart failure clinic. Patients were diagnosed between January 1, 2011 and December 31, 2019 and followed for an additional year to December 21, 2020. A set of baseline characteristics including age at time of diagnosis, sex (as assigned at birth), race, body mass index (BMI), health insurance status, Distressed Communities Index (DCI) score, and health comorbidities as identified by *International Classification of Diseases, Ninth Revision* (*ICD-9*) and *Tenth Revision* (*ICD-10*) codes as billing diagnoses or listed in Past Medical History or Problem List on index date – namely hypertension, hyperlipidemia, diabetes, heart failure, history of stroke or transient ischemic attack, vascular disease, chronic kidney disease, dialysis dependence, and chronic obstructive pulmonary disease – were gathered. A second set of characteristics specific to diagnosis of AF and treatment was collected including date of AF diagnosis, site of AF diagnosis (IM/PC, general cardiology, or heart failure clinic), attempt at cardioversion, anticoagulant prescription and anti-arrhythmic prescription, placement of order for referral to general cardiology, placement of order for referral to electrophysiology (EP), completion of visit with general cardiology, completion of visit with EP, and completion of AFCA.

The CHA_2_DS_2_-VASc score, a risk stratification calculator used to determine the one-year risk of a thromboembolic event in a non-anticoagulated patient with non-valvular AF, was calculated for each patient. Scores could range from 0 to 9; increasing scores correlated with increased risk of thromboembolism. A point was deducted for female sex, given its inclusion as a risk modifier in the original calculation. Patients with histories of use of warfarin that transitioned to DOACs were considered as DOAC recipients. BMI was calculated as mass divided by the square of the height (kg/m^2^). Self-reported residential zip codes were used to map patients with the DCI, an analytic tool designed to examine economic well-being across communities of the United States based on available Census Bureau data from 2010-2014. DCI scores based on patient’s reported zip codes could range from 0 (most prosperous) to 100 (most distressed).

An order for referral correlated to an order placed by a care provider in the EMR to recommend a patient to specialty care like cardiology or EP. A visit referred to the actual completion of an outpatient encounter with a specialist in cardiology or EP. It is possible that patients could be referred to a specialist without completing a visit and that patients could complete a visit to a specialist without a referral. A patient diagnosed in IM/PC could be referred to and/or visit with either general cardiology or EP. A patient diagnosed in general cardiology or heart failure clinic could be referred to and/or visit with EP. At Northwestern Medicine, a patient must be seen in the outpatient EP clinic setting prior to undergoing AFCA.

### Statistical Analysis

For analytical purposes, the study sample was limited to patients identified as non-Hispanic Black and non-Hispanic White, given the lower representation of other racial and ethnic groups. Baseline characteristics of the study participants were compared by sex utilizing a two-sample t-test for continuous variables and a Chi-square test for categorical variables. Separate logistic regression models were developed to evaluate sex disparities in each of the binary outcomes, including referral to general cardiology, visit with general cardiology, referral to EP, visit with EP, and AFCA completion. Odds ratios (ORs) and associated 95% confidence intervals (CIs) for women versus men were calculated from both unadjusted logistic regression models and models adjusted for a set of sociodemographic and clinical characteristics. In particular, models were adjusted for age at diagnosis, race/ethnicity, insurance type, body mass index, hyperlipidemia, hypertension, diabetes, heart failure, acute ischemic stroke, transient ischemic attack, coronary artery disease, peripheral artery disease, chronic kidney disease, dialysis, chronic obstructive pulmonary disease, obstructive sleep apnea, and DCI score.

Sensitivity analyses were performed to assess the impact of race on sex disparities, employing separate logistic regression models to examine the interaction of race and sex. ORs and 95% CIs were calculated by race, and p-values for the sex and race interaction were summarized. All analyses were conducted using SAS version 9.4 (Cary, NC).

## Results

This study’s overall cohort comprised of 5,445 non-Hispanic White and non-Hispanic Black patients originally diagnosed with AF in the outpatient setting. Of these 5,445 patients, 1,461 patients were diagnosed in IM/PC clinic, 3,711 patients in general cardiology clinic, and 273 patients in heart failure clinic (Figure 1). Among all 5,445 patients, 528 patients were referred to EP, and 1,815 patients visited directly with EP. Out of the 1,461 patients diagnosed in IM/PC clinic, 498 were referred to general cardiology clinic, and 695 visited with general cardiology.

**Figure 1.**
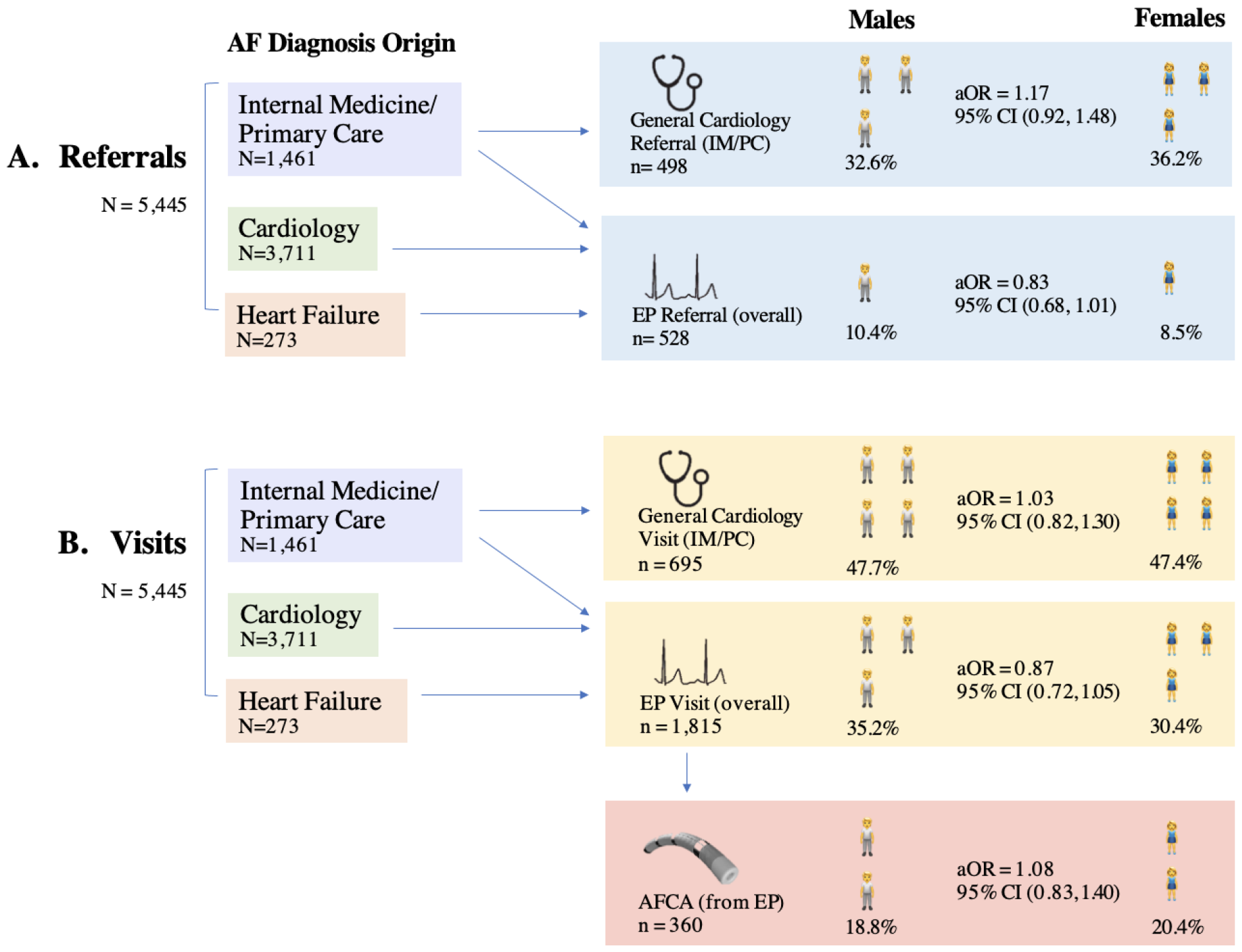
Pathway from AF to AFCA. **A**. Patients diagnosed in IM/PC clinic could be referred to general cardiology clinic. Patients diagnosed in either IM/PC, general cardiology, or heart failure clinic could be referred to EP. **B**. Patients diagnosed in IM/PC clinic could visit with general cardiology. Patients diagnosed in either IM/PC, general cardiology, or heart failure clinic could visit with EP. A subset of patients who visited with EP ultimately pursued AFCA. aOR indicates an adjusted odds ratio.

Of the overall 5,445 patients diagnosed with AF, 2,108 (38.7%) were female and 3,337 (61.3%) were male (Table 1). Women, on average, were older, had lower mean BMI, had higher CHA_2_DS_2_-VASc scores, and had higher DCI scores. A greater proportion of women were non-Hispanic Black in comparison to men. Women were more likely to receive Medicare, while men were more likely to hold private insurance. With respect to cardiovascular comorbidity profile, women were more likely to present with hypertension, transient ischemic attack or stroke, and chronic obstructive pulmonary disease, while men were more likely to have hyperlipidemia, vascular disease, chronic kidney disease, and obstructive sleep apnea. There was no significant difference in use of anticoagulation between men and women. Women were less likely to be placed on anti-arrhythmic agents than men. Lower usage of anti-arrhythmic agents by women was driven by differential rates of amiodarone and dofetilide prescription. More men were diagnosed in general cardiology and heart failure clinics, while more women were diagnosed in IM/PC clinic.

**Table 1:**
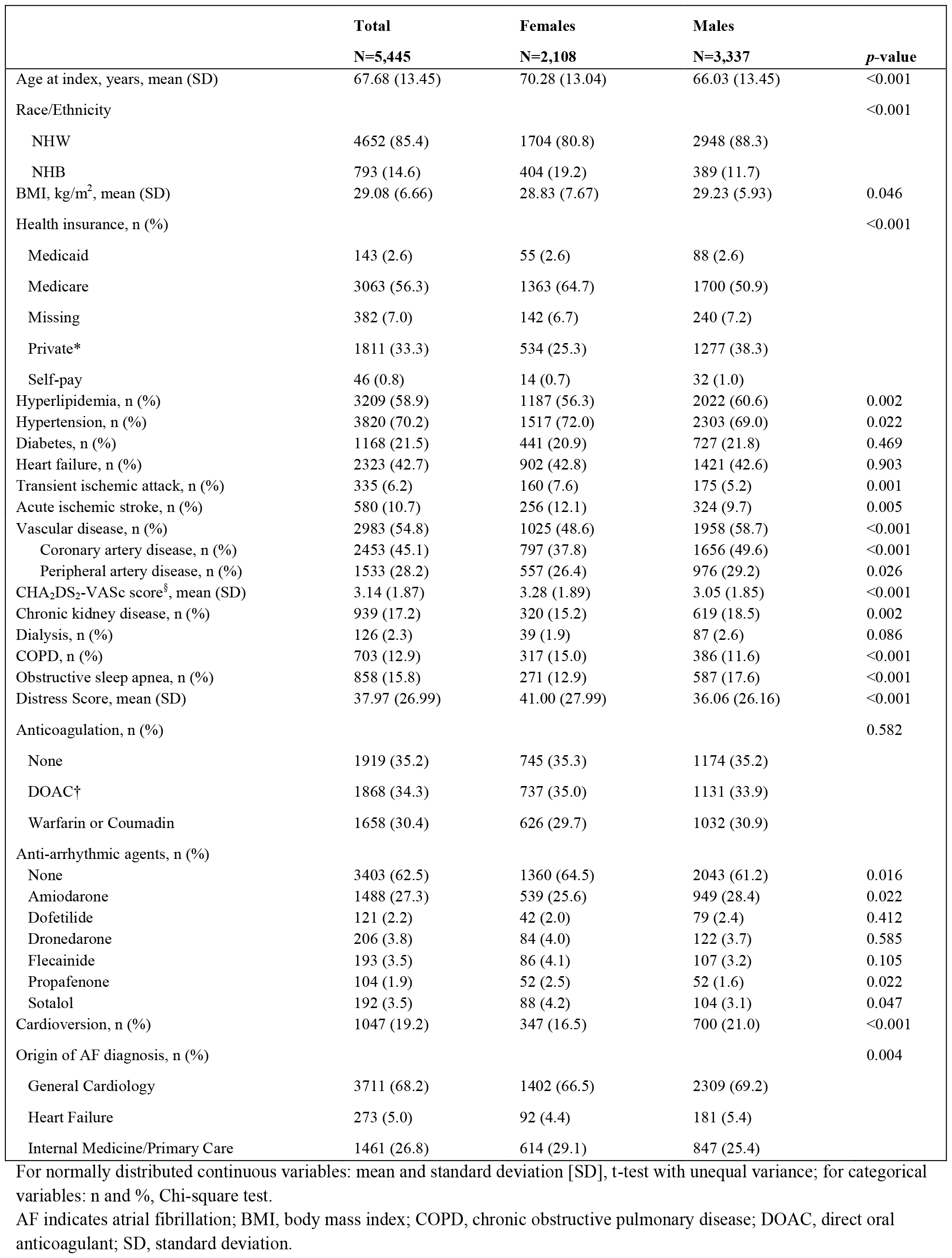
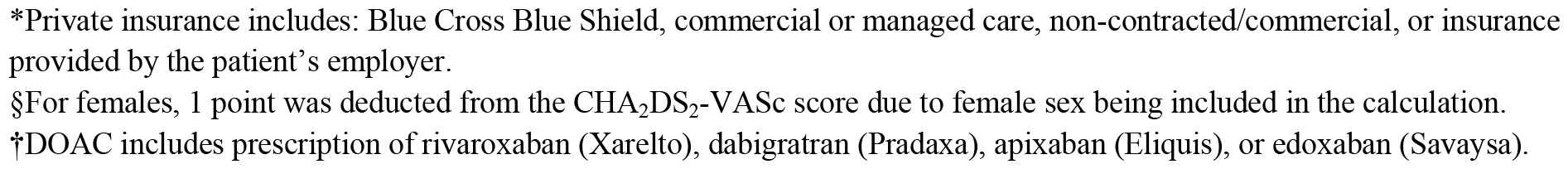
Baseline characteristics of the study population by sex.

|  | **Total** | **Females** | **Males** |  |
| --- | --- | --- | --- | --- |
|  | **N=5,445** | **N=2,108** | **N=3,337** | **p-value** |
| Age at index, years, mean (SD) | 67.68 (13.45) | 70.28 (13.04) | 66.03 (13.45) | <0.001 |
| Race/Ethnicity |  |  |  | <0.001 |
| NHW | 4652 (85.4) | 1704 (80.8) | 2948 (88.3) |  |
| NHB | 793 (14.6) | 404 (19.2) | 389 (11.7) |  |
| BMI, kg/m<sup>2</sup>, mean (SD) | 29.08 (6.66) | 28.83 (7.67) | 29.23 (5.93) | 0.046 |
| Health insurance, n (%) |  |  |  | <0.001 |
| Medicaid | 143 (2.6) | 55 (2.6) | 88 (2.6) |  |
| Medicare | 3063 (56.3) | 1363 (64.7) | 1700 (50.9) |  |
| Missing | 382 (7.0) | 142 (6.7) | 240 (7.2) |  |
| Private* | 1811 (33.3) | 534 (25.3) | 1277 (38.3) |  |
| Self-pay | 46 (0.8) | 14 (0.7) | 32 (1.0) |  |
| Hyperlipidemia, n (%) | 3209 (58.9) | 1187 (56.3) | 2022 (60.6) | 0.002 |
| Hypertension, n (%) | 3820 (70.2) | 1517 (72.0) | 2303 (69.0) | 0.022 |
| Diabetes, n (%) | 1168 (21.5) | 441 (20.9) | 727 (21.8) | 0.469 |
| Heart failure, n (%) | 2323 (42.7) | 902 (42.8) | 1421 (42.6) | 0.903 |
| Transient ischemic attack, n (%) | 335 (6.2) | 160 (7.6) | 175 (5.2) | 0.001 |
| Acute ischemic stroke, n (%) | 580 (10.7) | 256 (12.1) | 324 (9.7) | 0.005 |
| Vascular disease, n (%) | 2983 (54.8) | 1025 (48.6) | 1958 (58.7) | <0.001 |
| Coronary artery disease, n (%) | 2453 (45.1) | 797 (37.8) | 1656 (49.6) | <0.001 |
| Peripheral artery disease, n (%) | 1533 (28.2) | 557 (26.4) | 976 (29.2) | 0.026 |
| CHA<sub>2</sub>DS<sub>2</sub>-VASc score<sup>s</sup>, mean (SD) | 3.14 (1.87) | 3.28 (1.89) | 3.05 (1.85) | <0.001 |
| Chronic kidney disease, n (%) | 939 (17.2) | 320 (15.2) | 619 (18.5) | 0.002 |
| Dialysis, n (%) | 126 (2.3) | 39 (1.9) | 87 (2.6) | 0.086 |
| COPD, n (%) | 703 (12.9) | 317 (15.0) | 386 (11.6) | <0.001 |
| Obstructive sleep apnea, n (%) | 858 (15.8) | 271 (12.9) | 587 (17.6) | <0.001 |
| Distress Score, mean (SD) | 37.97 (26.99) | 41.00 (27.99) | 36.06 (26.16) | <0.001 |
| Anticoagulation, n (%) |  |  |  | 0.582 |
| None | 1919 (35.2) | 745 (35.3) | 1174 (35.2) |  |
| DOAC† | 1868 (34.3) | 737 (35.0) | 1131 (33.9) |  |
| Warfarin or Coumadin | 1658 (30.4) | 626 (29.7) | 1032 (30.9) |  |
| Anti-arrhythmic agents, n (%) |  |  |  |  |
| None | 3403 (62.5) | 1360 (64.5) | 2043 (61.2) | 0.016 |
| Amiodarone | 1488 (27.3) | 539 (25.6) | 949 (28.4) | 0.022 |
| Dofetilide | 121 (2.2) | 42 (2.0) | 79 (2.4) | 0.412 |
| Dronedarone | 206 (3.8) | 84 (4.0) | 122 (3.7) | 0.585 |
| Flecainide | 193 (3.5) | 86 (4.1) | 107 (3.2) | 0.105 |
| Propafenone | 104 (1.9) | 52 (2.5) | 52 (1.6) | 0.022 |
| Sotalol | 192 (3.5) | 88 (4.2) | 104 (3.1) | 0.047 |
| Cardioversion, n (%) | 1047 (19.2) | 347 (16.5) | 700 (21.0) | <0.001 |
| Origin of AF diagnosis, n (%) |  |  |  | 0.004 |
| General Cardiology | 3711 (68.2) | 1402 (66.5) | 2309 (69.2) |  |
| Heart Failure | 273 (5.0) | 92 (4.4) | 181 (5.4) |  |
| Internal Medicine/Primary Care | 1461 (26.8) | 614 (29.1) | 847 (25.4) |  |

For normally distributed continuous variables: mean and standard deviation [SD], t-test with unequal variance; for categorical variables: n and %, Chi-square test.
AF indicates atrial fibrillation; BMI, body mass index; COPD, chronic obstructive pulmonary disease; DOAC, direct oral anticoagulant; SD, standard deviation.
\*Private insurance includes: Blue Cross Blue Shield, commercial or managed care, non-contracted/commercial, or insurance provided by the patient's employer.
§For females, 1 point was deducted from the CHA<sub>2</sub>DS<sub>2</sub>-VASc score due to female sex being included in the calculation.
†DOAC includes prescription of rivaroxaban (Xarelto), dabigatran (Pradaxa), apixaban (Eliquis), or edoxaban (Savaysa).

In examining referral patterns to specialists for the management of AF among IM/PC patients, no significant differences were found in odds of referral to (adjusted OR [aOR], 1.17 [0.92-1.48], P=0.20) or visits with (aOR, 1.03 [0.82-1.30], P=0.79) general cardiologists between women and men (Table 2). Furthermore, in the overall cohort, there were no significant differences in the rates of referral to (aOR, 0.83 [0.68-1.01], P=0.06) or visits with (aOR, 0.87 [0.72-1.05], P=0.15) EP between women and men. Once seen by an electrophysiologist, there was no difference in the rates of proceeding to AFCA between women and men (aOR, 1.08 [0.83-1.40], P=0.56).

**Table 2:** Sex differences in referrals, visits, and catheter ablation for atrial fibrillation.

| Outcomes | Overall<br>n/N (%) | Males<br>n/N (%) | Females<br>n/N (%) | Unadjusted |  | Adjusted |  |
| --- | --- | --- | --- | --- | --- | --- | --- |
|  |  |  |  | Females vs.<br>Males OR (95%<br>CI) | p- value | Females vs.<br>Males OR (95%<br>CI) | p- value |
| Referral to General Cardiology (among IM/PC patients) | 498/1461<br>(34.1%) | 276/847<br>(32.6%) | 222/614<br>(36.2%) | 1.17 (0.94, 1.46) | 0.15 | 1.17 (0.92, 1.48) | 0.20 |
| Visit with General Cardiology (among IM/PC patients) | 695/1461<br>(47.6%) | 404/847<br>(47.7%) | 291/614<br>(47.4%) | 0.99 (0.80, 1.22) | 0.91 | 1.03 (0.82, 1.30) | 0.79 |
| Referral to EP | 528/5445<br>(9.7%) | 348/3337<br>(10.4%) | 180/2108<br>(8.5%) | 0.80 (0.66, 0.97) | 0.02 | 0.83 (0.68, 1.01) | 0.06 |
| Visit with EP | 1815/5445<br>(33.3%) | 1175/3337<br>(35.2%) | 640/2108<br>(30.4%) | 0.80 (0.71, 0.90) | <.001 | 0.87 (0.72, 1.05) | 0.15 |
| AFCA (among patients who visited with EP) | 360/1815<br>(19.8%) | 120/640<br>(18.8%) | 240/1175<br>(20.4%) | 0.90 (0.70, 1.15) | 0.39 | 1.08 (0.83, 1.40) | 0.56 |

AFCA indicates catheter ablation for atrial fibrillation; CI, confidence interval; EP, Electrophysiology outpatient clinic; IM/PC, Internal Medicine/Primary Care outpatient clinic; OR, odds ratio.
\*Adjusted for age at diagnosis, race/ethnicity, insurance type, body mass index, hyperlipidemia, hypertension, diabetes, heart failure, acute ischemic stroke, transient ischemic attack, coronary artery disease, peripheral artery disease, chronic kidney disease, dialysis, chronic obstructive pulmonary disease, obstructive sleep apnea, and Distressed Communities Index.

Sensitivity analyses were performed to evaluate synergistic effects of sex and race on the studied outcomes by including the interaction of sex and race in the logistic regression model. Across all outcomes, no statistically significant interactions between sex and race were observed (Table 3). Logistic regression models evaluating sex disparities separately among the two race groups yielded similar results to the models analyzing the overall cohort and were largely statistically insignificant except for the outcome of visits with EP. In both race groups, the unadjusted analysis showed that women were less likely to visit with EP than men; these sex disparities were statistically significant among non-Hispanic Black patients but not the non-Hispanic White patients and were statistically insignificant after adjusting for sociodemographic and clinical characteristics.

**Table 3.** Sex differences by race/ethnicity.

| **Outcomes** | **Females vs. Males**<br>**OR (95% CI)** | **Non-Hispanic Black** | **Non-Hispanic White** | **P-value for Sex and Race/ethnicity interaction** |
| --- | --- | --- | --- | --- |
| Referral to General Cardiology (among IM/PC patients), N=1461 | Unadjusted | 1.12 (0.69, 1.82) | 1.13 (0.88, 1.45) | 0.96 |
|  | Adjusted * | 1.17 (0.71, 1.93) | 1.17 (0.90, 1.53) | 0.99 |
| Visit with General Cardiology (among IM/PC patients), N=1461 | Unadjusted | 0.92 (0.56, 1.48) | 1.00 (0.79, 1.26) | 0.75 |
|  | Adjusted * | 0.91 (0.5, 1.49) | 1.06 (0.83, 1.37) | 0.57 |
| Visit with EP, N=5445 | Unadjusted | 0.77 (0.60, 0.98) | 0.88 (0.70, 1.10) | 0.42 |
|  | Adjusted * | 0.79 (0.61, 1.04) | 0.94 (0.73, 1.20) | 0.34 |
| AFCA (among patients who visited with EP), N=1815 | Unadjusted | 0.69 (0.32, 1.49) | 0.95 (0.74, 1.23) | 0.44 |
|  | Adjusted * | 0.87 (0.40, 1.92) | 1.11 (0.84, 1.45) | 0.58 |

AFCA indicates catheter ablation for atrial fibrillation; CI, confidence interval; EP, Electrophysiology outpatient clinic; IM/PC, Internal Medicine/Primary Care outpatient clinic; OR, odds ratio.
\*Adjusted for age at diagnosis, insurance type, body mass index, hyperlipidemia, hypertension, diabetes, heart failure, acute ischemic stroke, transient ischemic attack, coronary artery disease, peripheral artery disease, chronic kidney disease, dialysis, chronic obstructive pulmonary disease, obstructive sleep apnea, and Distressed Communities Index.

## Discussion

We did not observe a sex disparity in referral patterns to specialists nor in the utilization of AFCA within our single integrated health system. Among patients who visited with EP, women were equally likely to undergo AFCA compared with their male counterparts.

Prior literature has demonstrated that women are less likely to undergo AFCA compared to men.^1,12, 14, 20-23^ Lower utilization of AFCA for women is thought to be related to a few factors. Studies have found that women undergoing AFCA are generally older, have a longer history of AF than men, and have a higher prevalence of hypertension.^24, 25^ Older age and related comorbidities may confer greater procedural risk and drive physicians to restrict AFCA performance. A consensus statement from the European Heart Rhythm Association posits that women undergo AFCA less frequently due to patient preference and hesitation regarding invasive treatment.^26^ Some studies demonstrate poorer results after AFCA in women, likely related to women having AF progression and complex electrophysiological anatomy in the setting of longer diagnosis-to-procedure time.^27-29^ Our study represents a departure from prior literature in that we observed that, among patients who visited with EP, women were just as likely to undergo AFCA as men. Additionally, we found that men and women were equally likely to be referred to general cardiology and EP, suggesting that differences in referral patterns between men and women do not explain the historically demonstrated sex disparity in AFCA utilization.

This study also found that the number of patients who visited with general cardiology or EP was greater than the number referred to these clinics. Among all 5,445 patients diagnosed in the outpatient setting, 528 patients were referred to EP, while a larger 1,815 patients visited directly with EP. Additionally, though only 498 were referred to general cardiology clinic, 695 ultimately visited with general cardiology. Such differences between the number of patients referred to specialty services and the number of patients who ultimately visited with these specialists highlight that patients are not dependent on referral orders to visit with specialists. At this health system, patients can self-refer to specialists, a common occurrence especially for patients lacking establishment with primary care. Additionally, patients may have appointments directly made with specialty services after emergency department visits or inpatient stays. Though fewer referral orders were placed than actual visits completed, no significant sex disparities were observed in either referral orders or specialty visits, illustrating that bypass of referrals to visit specialists did not confound this study’s examination of referral patterns.

The present study also highlights a unique interplay between sex, race, and visits with specialists, namely EP. We found in the unadjusted analysis that among non-Hispanic Black patients, women were less likely than men to visit with EP. This finding builds on prior work by Duke et al. who found that among patients who visited with EP, non-Hispanic Black patients were less likely to undergo AFCA compared with non-Hispanic White patients.^19^ While our study did not find statistically significant differences in AFCA utilization between women and men among non-Hispanic Black and non-Hispanic White patients, our study does note a connection between sex and race that affects a non-Hispanic Black female patient’s access to specialist care compared to her male counterparts. This statistically significant difference in specialist visits observed in our study disappears once adjusted for socioeconomic and clinical characteristics, suggesting a possible interaction between socioeconomic status and referral patterns for AFCA. Further characterization of this relationship will allow us to better understand the interactions of sex, race, and socioeconomic status and its effects on procedural management of AF.

This study is limited by its single-center, retrospective nature and reliance on EMR categorization of sex. This study was not designed to determine sex differences in outcome after treatment. Data was also not collected on the lengths of time between initial diagnosis of AF, visits with specialists, and completion of AFCA. Further characterization of the sex disparity in AFCA utilization could lie in the examination of sex-based differences in post-operative outcomes, complication rates, and diagnosis-to-procedure time.^16-18, 27-29^

## Conclusions

Our study demonstrates no significant differences in referral patterns to specialists or in rates of AFCA between individuals of different sex who receive care from a single integrated health system. Encouraging equitable referral to specialists and access to AF ablation is essential in ensuring appropriate care for all patients.

## Data Availability

The data that support the findings of this study are available from the corresponding author upon reasonable request.

## Acknowledgments

All authors take responsibility for the decision to submit the article for publication. Drs. Gangadharan, Muhammad, Knight, and Passman and Ms. Song had complete access to the data analyzed in this study and take full responsibility for the integrity of the data and its analysis.

